# Evaluating Neurotrophins and Irisin in a Lifestyle Intervention Study for Dementia Risk Reduction

**DOI:** 10.1101/2025.09.08.25335367

**Authors:** Rhys Houston, Thomas Pace, Sophie C. Andrews, Bonnie L. Quigley

## Abstract

**Background:** The rising prevalence of Alzheimer’s disease (AD) and related neurodegenerative disorders highlights the need for non-pharmacological strategies to preserve cognitive health. Brain-derived neurotrophic factor (BDNF), vascular endothelial growth factor (VEGF), and irisin are biomarkers associated with cognitive resilience and metabolic health.

**Objective:** To assess the impact of a multimodal lifestyle intervention on serum BDNF, VEGF, and irisin levels in an aging population, aiming to explore how modifiable lifestyle factors influence biomarkers linked to neurodegeneration risk.

**Methods:** In a randomised controlled trial, participants were assigned to a 12-week lifestyle intervention group (n=42, 32 female; mean age 67.1±7.6 years) or a control group (n=41, 34 female; mean age 62.7±8.6 years). The intervention included structured physical exercise, dietary modifications, sleep hygiene, and mindfulness practices. Serum BDNF, VEGF, and irisin levels were measured at baseline (T1), post-intervention (T2), and six months post-intervention (T3). Linear mixed models assessed biomarker changes over time and between groups.

**Results:** Irisin levels increased significantly post-intervention (T2) in the intervention group (p < 0.001) in comparison to the control group and were maintained at T3 (p = 0.003). The intervention group also exhibited a significant attenuation of BDNF decline at T3 compared to the control group (p = 0.007). VEGF levels remained stable across all time points (p > 0.05). Collectively, these changes suggest that the intervention resulted in positive metabolic and neuroprotective effects.

**Conclusion:** A structured lifestyle intervention can significantly elevate BDNF and irisin levels, supporting non-pharmacological strategies to enhance brain health and potentially reduce neurodegeneration risk in older adults.

## Introduction

Alzheimer’s disease (AD) is the leading cause of dementia in older adults, with prevalence expected to triple by 2050 (1). AD is characterized by progressive cognitive decline, driven by the accumulation of amyloid-beta (Aβ) plaques and neurofibrillary tangles (NFTs) in the brain (2, 3). These pathological proteins disrupt neuronal communication, impair synaptic function, and contribute to hippocampal atrophy, a hallmark of memory loss and cognitive dysfunction (4, 5). Given the limited efficacy of pharmacological treatments, there is growing interest in non-pharmacological strategies to preserve brain health and delay neurodegeneration.

Emerging research highlights the roles of three key proteins in supporting cognitive resilience: brain-derived neurotrophic factor (BDNF), vascular endothelial growth factor (VEGF), and irisin. BDNF, highly expressed in the hippocampus and cerebral cortex, promotes neurogenesis, synaptic plasticity, and neuronal survival (6, 7). VEGF supports cerebrovascular health by facilitating angiogenesis, ensuring adequate oxygen and nutrient delivery to neurons (8). Irisin, a myokine released during exercise (9), crosses the blood-brain barrier and enhances BDNF expression, creating a feedback loop that fosters neuroplasticity (10). Together, these proteins represent critical components in the maintenance of cognitive function (11).

While exercise has been shown to elevate BDNF and irisin levels and support VEGF-mediated vascular health, few studies have examined the combined effects of multimodal lifestyle interventions on all three (12). Specifically, the influence of sleep hygiene, mindfulness practices, and dietary modifications, in conjunction with physical activity, remains underexplored (13). Understanding how these modifiable factors impact biomarker levels could inform preventive strategies aimed at reducing neurodegeneration risk in aging populations.

This study aimed to investigate the effects of a 12-week comprehensive lifestyle intervention which included exercise, diet, sleep hygiene, and mindfulness (13) on serum levels of BDNF, VEGF, and irisin. We hypothesized that such interventions would significantly elevate the circulating levels of all three proteins, supporting neuroplasticity and metabolic activity, with potential implications for reducing the risk of neurodegeneration in older adults.

## Methods

### Study design

This study is a sub-analysis from the Lifestyle Intervention Study for Dementia Risk Reduction (LEISURE) trail conducted at the University of the Sunshine Coast’s Thompson Institute (ACTRN ID ACTRN12620000054910, UniSC Ethics Approval: A19130)(13). The 24-month, parallel-group randomised control trial included independently living adults aged 50-85 years. Inclusion criteria required normal or correct-to-normal vision and absence of dementia or mild cognitive impairment, as confirmed by the Montreal Cognitive Assessment (MoCA) and psychological assessments. All participants provided informed consent. Participants underwent baseline assessments and received a personalised Dementia Risk Report (DRR) and were then randomly assigned to either a 12-week multimodal lifestyle intervention group (LIFE) or a health-information-only control group (TAU). Primary outcomes of this study will be reported elsewhere. Study timepoints included assessment at baseline (T1), post-intervention (T2), and six months post-intervention (T3).

### Intervention Group

The 12-week LIFE intervention included four components: 1) Exercise: Three weekly 60-minute sessions of aerobic, strength, and balance training, conducted under certified physiologist supervision in a gym setting; 2) Diet: Six biweekly group sessions led by a qualified dietitian, focusing on adherence to a modified Mediterranean diet using SMART goals; 3) Mindfulness: Six biweekly 90-minute sessions led by a trained mindfulness instructor, with daily home practice (starting at 20 minutes, increasing to 30 minutes); 4) Sleep hygiene: Six biweekly 60-minute sessions facilitated by a behavioural sleep psychologist, incorporating education on circadian rhythms and sleep disorders, with personalised goal setting.

### Control Group

Control participants received their DRR but no additional intervention. They were encouraged to consult regular healthcare providers. After T2 assessments, they gained online access to general health information.

### Biological Sample Processing and Assays

Fasting blood samples were collected in serum SST tubes, allowed to clot for 30 minutes, and centrifuged at 3500 rpm (2456 x g) at 4°C for 15 minutes. Serum aliquots were stored at -80°C until analysis. BDNF and VEGF levels were quantified in duplicate using ProcartaPlex Simplex beads on a Luminex 200 multiplex immunoassay system (ThermoFisher Scientific), following manufacturers protocols. Irisin levels were measured in duplicate using a commercial ELISA kit (Phoenix Pharmaceuticals, USA), following manufacturers protocols. From the LEISURE study participant pool, 83 baseline samples (T1) (66 female, 17 male), 59 post-intervention (T2) samples, and 54 six-month follow-up (T3) samples were available for analysis.

### Statistical Analysis

Data analysis was performed in RStudio (version 2024.4.2.764) (14). Demographic variables between groups were assessed using Chi squared, independent samples t-tests and Wilcoxon rank-sum tests as appropriate. Linear mixed models (LMMs), implemented via the lme4 and lmerTest packages (15, 16), assessed fixed effects (intervention, time, age) and random effects (participants). Model comparisons were performed using ANOVA, considering Akaike Information Criterion (AIC), Bayesian Information Criterion (BIC), and chi-square values. Biomarker data were transformed to reflect changes from baseline, standardising comparisons across participants.

## Results

### Study Population

Demographic comparisons at baseline showed no significant differences between the intervention (n = 42) and control (n = 41) groups in sex distribution (p = 0.835) or education level (p = 0.070) (Table 1). However, the intervention group ended up being an average of five years older than the control group (67.1 ± 7.6 years vs. 62.7 ± 8.6 years, *t*(79.1) = 2.49, p = 0.015). This age difference had the follow-on effect of significant average baseline differences between groups in BDNF (p = 0.040) and VEGF (p = 0.002) group levels (Table 1). The age difference between groups did not impact irisin group levels (p = 0.109) (Table 1).

**Table 1:** Descriptive statistics and baseline comparisons between control and intervention groups.

| Characteristic | Control Group<br>(n = 41) | Intervention<br>Group (n = 42) | Statistic | P Value |
| --- | --- | --- | --- | --- |
| Sex | 34 F / 7 M | 32 F / 10 M | $\chi^2(1) = 0.04$ | 0.835 |
| Age | 62.7 ± 8.6 | 67.1 ± 7.6 | $t(79.124) = 2.49$ | **0.015** |
| Education | 15.4 ± 3.02 | 15.0 ± 4.70 | $W = 1034$ | 0.070 |
| BDNF (baseline, pg/ml) | 145.0 ± 136.0 | 83.3 ± 58.4 | $W = 1087$ | **0.040** |
| VEGF (baseline, pg/ml) | 368.0 ± 221.0 | 219.0 ± 171.0 | $W = 1204$ | **0.002** |
| Irisin (baseline, ng/ml) | 6.19 ± 3.61 | 5.71 ± 4.1 | $W = 1037$ | 0.109 |

### Biomarker Changes Over Time

To mitigate the average baseline differences in BDNF and VEGF levels between the intervention and control groups, all the biomarker data was transformed to change values. Change values for each biomarker which were calculated by subtracting the baseline levels from subsequent timepoint levels (similar to (17)). This set each participant’s baseline value to “zero” and converted subsequent timepoint values to the change in biomarker level from baseline. Models then included age as a fixed effect to account for any remaining impact of the group-level age differences on biomarkers changes over time.

### BDNF

Linear mixed model analysis revealed a significant interaction effect at T3 between the intervention and control groups (p = 0.019). While the control group exhibited a significant decline in BDNF levels from T1 to T3 (p = 0.024), the intervention group maintained stable BDNF levels (p = 0.789). Between-group comparisons at T3 demonstrated significantly higher BDNF levels in the intervention group compared to the control group (p = 0.007), suggesting that the intervention effectively attenuated age-related declines in BDNF over time (Figure 1A).

**Figure 1:**
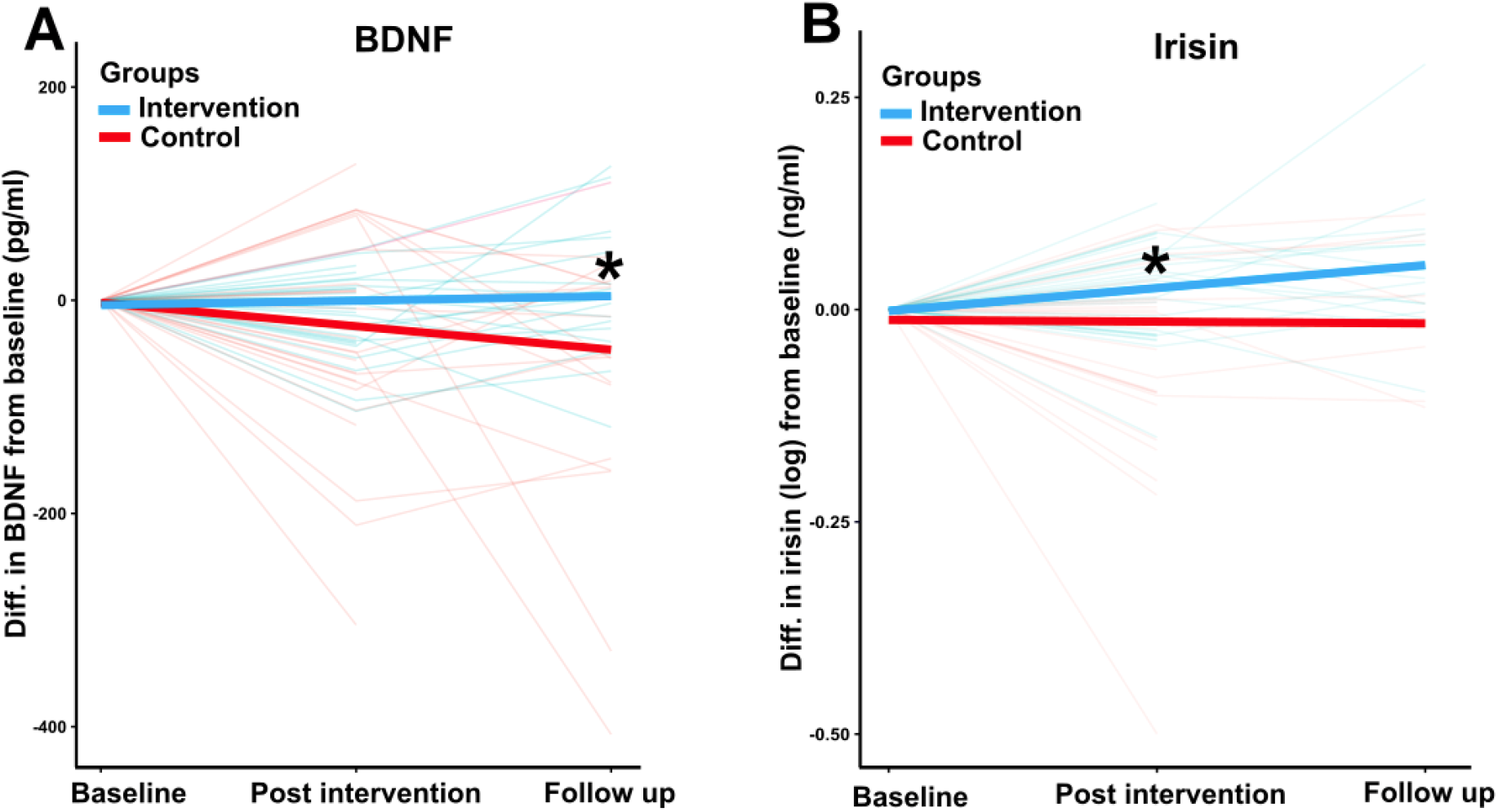
Linear Mixed Model changes in biomarker levels over time for the intervention and control groups. This line plot illustrates the results from the LMM with the best model fit showing changes in biomarker levels (pg/ml for BDNF, ng/ml for irisin) across the time points (baseline, post-intervention, follow-up) for both the intervention group (blue lines) and the control group (red lines). The solid line represents the mean change in biomarker levels over time, and the lighter line represent individual participants trajectories. Asterisks denote points of significant differences.

### VEGF

VEGF levels remained stable across all time points, with no significant differences observed within or between groups (p > 0.05). The best-fit LMM suggested changes in VEGF levels were independent of the intervention and participant age. These results indicate that the multimodal lifestyle intervention had limited effects on vascular remodelling within the study period.

### Irisin

Irisin levels showed a significant increase at T2 in the intervention group compared to the control group (p < 0.001). The intervention group also maintained elevated irisin levels at T3 (p = 0.003), whereas the control group experienced a decline post-intervention. These findings indicate that the intervention had an immediate and sustained positive effect on irisin levels (Figure 1B).

## Discussion

The aim of this study was to investigate the effects of a multimodal dementia risk lifestyle intervention on three key proteins related to AD: BDNF, VEGF, and irisin. Significant time-dependent increases in BDNF and irisin levels were observed in the intervention group, while VEGF levels remained largely unchanged.

Data from healthy aging participants (n = 83) were analysed, with participants randomised into intervention (n = 42) and control groups (n = 41). The LEISURE study (13) recruited predominately women (n = 66), consistent with trends showing women are more likely to volunteer for health-related research (18). Age differences between groups were accounted for by transforming the data to change from baseline levels and including age as a fixed effect in the linear mixed models, minimising potential confounding effects.

The increase in BDNF aligns with research linking BDNF to neuroplasticity and cognitive function (19-21). The multimodal intervention likely promoted neurobiological processes enhancing BDNF expression. In contrast, VEGF may require specific vascular stimuli, such as hypoxia or sustained high-intensity exercise, which were not central components of the LEISURE intervention (22).

The observed relationship between BDNF and irisin supports recent findings that exercise-induced irisin elevates BDNF expression (23). Irisin can cross the blood-brain barrier, promoting neuroplasticity and cognitive performance (24). The concurrent increase in both proteins suggest synergistic effects of the intervention on neuronal health (23, 24).

Despite previous studies reporting VEGF’s role in enhancing neuroplasticity alongside BDNF, VEGF levels in this study did not significantly change (25-27). This discrepancy may be due to the intensity and timing of exercise sessions, as VEGF responses to exercise are transient and highly dependent on exercise type and timing of measurement (22). Additionally, participants’ higher baseline fitness levels may have influenced VEGF regulation, contributing to the absence of significant VEGF changes (28).

The longitudinal design of the LEISURE study enabled the detection of sustained BDNF and irisin elevations up to six months post-intervention. This findings aligns with meta-analyses showing that aerobic exercise can induce prolonged increases in resting BDNF levels, likely through mechanisms such as enhanced cerebral blood flow and neurogenesis (29). The sustained elevation of BDNF and irisin emphasises the potential long-term neuroprotective effects of multimodal lifestyle interventions in healthy aging populations.

To conclude, this study highlights the potential of multimodal lifestyle interventions-including exercise, sleep hygiene, diet, and mindfulness in promoting biomarkers associated with neuroplasticity and cognitive health. The observed sustained increases in BDNF and irisin levels suggest that such interventions could have lasting neuroprotective effects in healthy aging populations. The lack of significant VEGF changes indicates that more intensive or specific vascular stimuli may be necessary to influence this factor. Future research should explore the dose-response relationship of exercise intensity and duration on VEGF levels, as well as the interplay between BDNF and irisin in mediating cognitive outcomes. Overall, these findings underscore the promise of holistic lifestyle approaches for mitigating dementia risk and supporting healthy brain aging.

## Data Availability

All data produced in the present study are available upon reasonable request to the authors.

## Supplementary Data

Available on request from author.

## Declaration of Conflicts of Interest

None.

## Funding Statement

No external funding.

## Notes

### Competing Interest Statement

The authors have declared no competing interest.

### Clinical Trial

Australian New Zealand Clinical Trials registry ACTRN12620000054910 (https://www.anzctr.org.au/Trial/Registration/TrialReview.aspx?id=379049)

### Clinical Protocols

https://pubmed.ncbi.nlm.nih.gov/37334601/

### Funding Statement

This study did not receive any funding.

### Author Declarations

Ethics committee/IRB of University of the Sunshine Coast gave ethical approval for this work (approval number A19130).

## References

1. Perveen A, Nephew, YL. Global, regional, and national burden of Alzheimer’s disease and other dementias, 1990 - 2019. Front Aging Neurosci. 2022.

2. Deture MA. DW. D. The neuropathological diagnosis of Alzheimer’s disease. Mol Degener. 2019;14.

3. Kapitein LC, T. H, Gouras G. Building the neuronal mictrotubule cytoskeleton. Cell Press. 2015;87:492–506.

4. Ashrafian H, Zadeh EH. RH. K. Review on Alzheimer’s disease: Inhibition of amyloid beta and tau tangle formation. Int J Biol Macromol. 2021;167:382–94.

5. Pini L, Pievani M, Bocchetta M, Altomare D, Bosco P, Cavedo E, et al. Brain atrophy in Alzheimer’s disease and aging. Aging Res Rev. 2016;30:25–48.

6. Park H, mm. P. Neurotrophin regulation of neural circuit development and function. Nat Revs Neurosci. 2012;14:7–23.

7. Bramham CR, E. M. BDNF function in adult synaptic plasticity: The synaptic consolidation hypothesis. Prog Neurobiol. 2012;76:609–42.

8. Perez-Gutierrez L. N. F. Biology and therapeutic targeting of vascular endothelial growth factor A. Nat Rev Mol Cell Bio. 2023;24:816–34.

9. Jin Y, Sumsuzzman D, Choi J, Kang H, Lee Sr, Y. H. Molecular and functional interaction of the myokine irisin with physical exercise and Alzheimer’s disease. Molecules. 2018;23.

10. Waseem R, Shamsi A, Mohammad T, Hassan MI, Kazim SN, *et. al*. FNDC5/Irisin: Physiology and Pathophysiology. Molecules. 2022;27(1118).

11. Reddy I, Yadav Y, CS. D. Cellular and molecular regulation of exercise - a neuronal perspective. Cell mol neurobiol. 2022;43:1551–71.

12. al. MKe. The Finnish Geriatric Intervention Study to Prevent Cognitive Impairment and Disability (FINGER): Study design and progress. Alzheimer’s & Dementia. 2013;9:657–65.

13. Treacy C, Levenstein JM, Jefferies A, Metse AP, Schaumberg MA, *et. al*. The LEISURE study: A longitudinal randomized controlled trial protocol for a multi-modal lifestyle intervention study to reduce dementia risk in healthy older adults. J Alzheimers Dis 2023;94:841–56.

14. Team P. RStudio: Integrated Development Environment for R. 2024.4.2.764 ed: Posit Software, PBC; 2024.

15. Bates D, Maechler M, Bolker B. S. W. Fitting linear Mixed-Effects Models Using lme4. J. Stat. Softw. 2015.

16. Kuznetsova A, Brockhodd B, Rune H. B C. lmerTest package: Tests in linear mixed effects models. J Stat Softw. 2017;82:1–26.

17. Zalta AK, Voigt RM, Stevens SK, Held P, Raeisi S, Boley RA, et al. Brain derived neurotrophic factor and treatment outcomes among veterans attending an intensive treatment program for posttraumatic stress disorder. J Psychiatr Res. 2024;173:1–5.

18. Garcia JS, Gil-Lacruz AI. M. G-L. The influence of gender equality on volunteering among european senior citizens. Voluntas. 2022;33:820–32.

19. Szuhany KL, Bugatti M. MW. O. A meta-analytic review of the effects of exercise on brain-derived neurotrophic factor. J Psychiatr Res. 2015;60:56–64.

20. Huang T, Larsen KT, Ried-Larsen M, Moller NC. LB. A. The effects of physical activity and exercise on brain-derived neurotrophic factor in healthy humans: A review. Scand J Med Sci Sports. 2014;24:1–10.

21. Vaynman S, Ying Z. F. G-P. Hippocampal BDNF mediates the efficacy of exercise on synaptic plasticity and cognition. EJN. 2004;20:2580–90.

22. Hoier B. Y. H. Exercise-induced capillary growth in human skeletal muscle and the dyamics of VEGF. Microcirculation. 2014;21:301–14.

23. Lourenco MV, Frozza RL, de Freitas G, Zhang H, Kincheski GC, *et. al*. Exercise-linked FNDC5/irisin rescues synaptic plasticity and memory defects in Alzheimer’s models. Nat Medicine. 2019;25:165–75.

24. Peng J. J. W. Effects of the FNDC5/Irisin on elderly dementia and cognitive impairment. Front Aging Neurosci. 2022;14.

25. Pickersgill J, Turco CV, Ramdeo K, Reshi RS, Foglia SD. AJ. N. The combined influences of exercise, diet and sleep on neuroplasticity. Front Psychol. 2022;13.

26. El-Sayes J, Harasym D, Turco CV, Locke MB. AJ. N. Exercise-induced neuroplasticity: A mechanism model and prospects for promoting plasticity. Neuroscientist. 2019;25:65–85.

27. Deyama S, Bang E, Kato T, Li XY. R. D. Neurotrophic and antidepressant actions of braindervied neurotrophic factor require vascular endothalial growth factor. Biol Psychiatry. 2019;86:143–52.

28. Wahl P, Zinner C, Achtzehn S, Behringer M. W. B. Effects of acid-base balance and high or low intensity exercise on VEGF and bFGF. Eur J Appl Physiol. 2011(111):1405–13.

29. Dinoff A HN, Swardfager W, Liu CS, Sherman C, Chan S, Lanctot KL. The effect of exercise on resting concentrations of peripheral brain-derived neurotrophic factor (BDNF): A metaanalysis. PLoS ONE. 2016;11.

